# Preparedness and Mitigation by projecting the risk against COVID-19 transmission using Machine Learning Techniques

**DOI:** 10.1101/2020.04.26.20080655

**Authors:** Akshay Kumar, Farhan Mohammad Khan, Rajiv Gupta, Harish Puppala

**Affiliations:** Birla Institute of Technology and Science, Pilani, Pilani Campus, Rajasthan, 333031, BML Munjal University, Gurugram

**Keywords:** COVID-19, Hotspots, Machine learning, Transmission, Lockdown

## Abstract

The outbreak of COVID-19 is first identified in China, which later spread to various parts of the globe and was pronounced pandemic by the World Health Organization (WHO). The disease of transmissible person-to-person pneumonia caused by the extreme acute respiratory coronavirus 2 syndrome (SARS-COV-2, also known as COVID-19), has sparked a global warning. Thermal screening, quarantining, and later lockdown were methods employed by various nations to contain the spread of the virus. Though exercising various possible plans to contain the spread help in mitigating the effect of COVID-19, projecting the rise and preparing to face the crisis would help in minimizing the effect. In the scenario, this study attempts to use Machine Learning tools to forecast the possible rise in the number of cases by considering the data of daily new cases. To capture the uncertainty, three different techniques: (i) Decision Tree algorithm, (ii) Support Vector Machine algorithm, and (iii) Gaussian process regression are used to project the data and capture the possible deviation. Based on the projection of new cases, recovered cases, deceased cases, medical facilities, population density, number of tests conducted, and facilities of services, are considered to define the criticality index (CI). CI is used to classify all the districts of the country in the regions of high risk, low risk, and moderate risk. An online dashpot is created, which updates the data on daily bases for the next four weeks. The prospective suggestions of this study would aid in planning the strategies to apply the lockdown/ any other plan for any country, which can take other parameters to define the CI.

## Introduction

The disease of transmissible person-to-person pneumonia caused by the extreme acute respiratory coronavirus 2 syndrome (SARS-COV-2, also known as COVID-19), has sparked a global warning The first case of COVID-19 is reported in India on 30^th^ January 2020. Starting from closing of the international border, travel, and entry restrictions, GoI imposed the trail lockdown (22^nd^ March), 21-day lockdown (from 24^th^ March-14^th^ April 2020), which is further extended by 19 days till May 3^rd^, with review on 20^th^ April 2020. A complete lockdown for a country like India, or for any other country, generates complexity. Not only laborers, farmers, and workers will lose daily earnings, but a major loss in the GDP of the country is unavoidable. Hence, various brainstorming sessions are being conducted by GoI to explore the possibility of opening the lockdown in a strategic manner. However, a small mistake may cause irreparable loss, and there is no going back, which is observed by other countries like China, Singapore, and others. Also, lockdown cannot be imposed for infinitely, and lifting it all together for the entire country is also not strategically correct, which may consequently resurge the COVID-19 cases and becomes uncontrollable with the existing resources. Delay in lifting the lockdown also may not be right considering the people's mental health and on the other systems like education, culture, and social.

## 2. Methodology

A prospective methodology is proposed to understand the spatial foot-print of COVID and to predict the possible spread along with analyzing the risk in a region. In this regard, a framework is designed to facilitate the decision-maker in forecasting the possible scenarios. The steps involved in the proposed approach are shown in Figure 1. From the flow chart presented below, it is apparent that the eight different parameters are considered in quantifying the risk associated with each of the regions using *Technique for Order of Preference by Similarity to Ideal Solution* (TOPSIS); a multi-criteria procedure.

**Figure 1.**
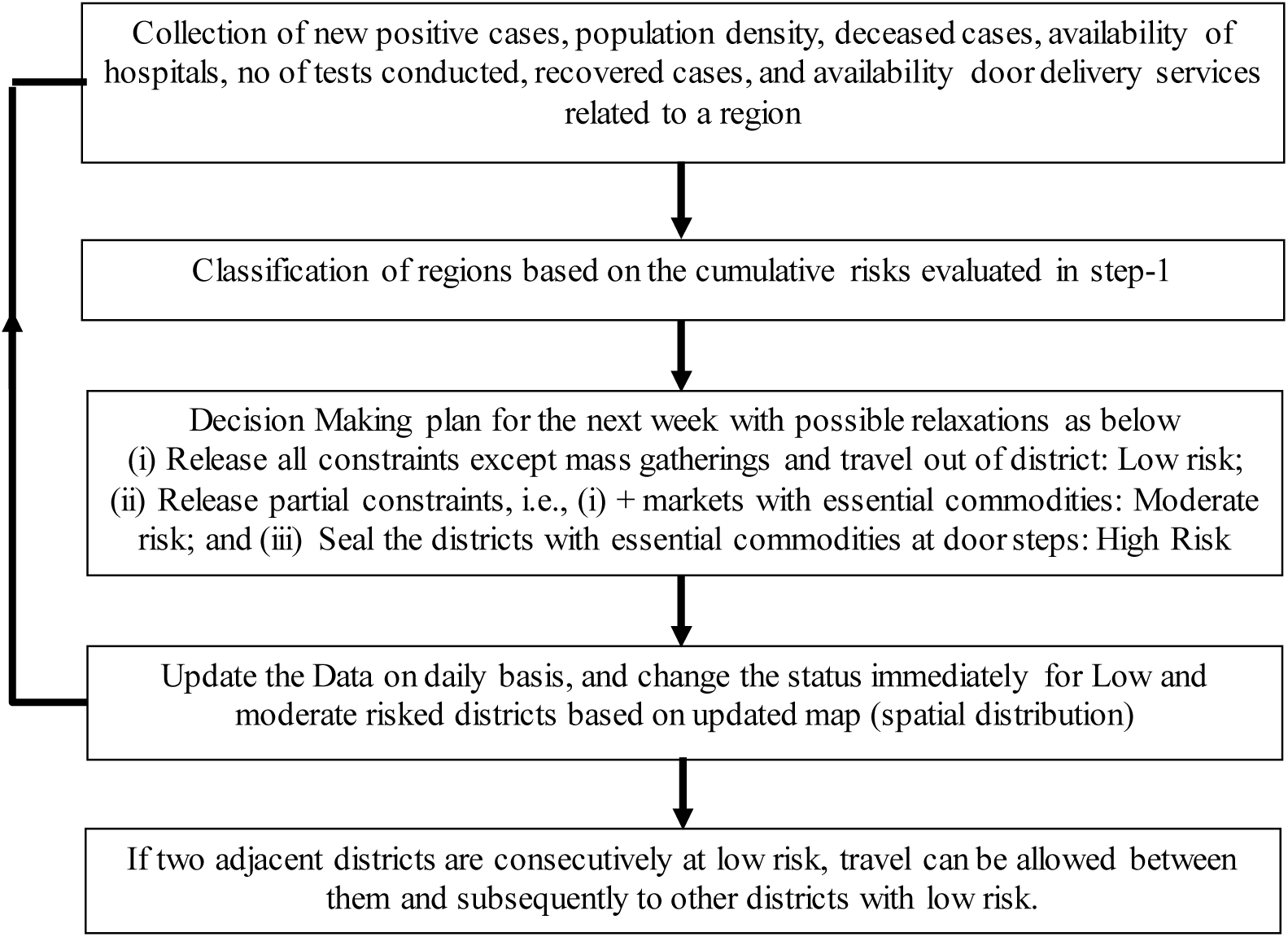
Algorithm of the proposed strategy

The different parameters considered are daily positive new cases, population, population density, deceased cases, availability of hospitals, no of tests conducted, recovered cases, and availability door delivery services. The considered positive cases define the intensity of COVID-19 in a region. Population data reflects the possible asymptomatic carriers. Population density, on the other hand, defines the risk of spread/community transmission. The number of deceased and recovered cases in the region determines the quality of health care facilities. The availability of hospital defines the strength to handle this detrimental situation. The risk associated with a region can be analyzed by considering the test reports. The availability of daily needs to the doorsteps is confined only to a few of the areas. These services help to minimize the mass gathering and mitigate the crowd at the retail units. The daily data corresponding to each of these variables is collected from various sources and is compiled in a repository.

Decision trees are easy to interpret, fast for fitting and predictions and, low on memory usage. It contains a coarse tree, medium tree, fine tree to increase the model flexibility with the maximum number of splits settings.

## 3. Analysis, Results and Discussion

The attribute related to the criteria discussed in the aforementioned sections is collected from 30^th^ Jan. Subsequently, the probable variation for the next four weeks with a time interval of one week is forecasted using the three different techniques, as discussed above. The forecasted attributes consequently help in estimating the cumulative score of the region, which is determined using *TOPSIS*, a Multi-criteria decision-making technique. The obtained cumulative score is introduced as the criticality index. Based on this distribution of criticality index, the regions can be classified into clusters of low risk, moderate risk, and high risk. Lockdown should be imposed in the area of high risk, whereas red zones can be identified in the regions of moderate-risk, and restriction to movement can be imposed. Lockdown in the region of low risk can be released with some precautionary measures. The probable variation of the criticality index for the next four weeks from 19^th^ April is mapped using the three different techniques. The variation obtained from TOPSIS using the data forecasted using DT, SVM, and GPR are shown in Figure 2, Figure 3, and Figure 4, respectively. For detailed understanding of DT, SVM and GPR refer the study of Zhang et. al., (2018), Pelckmans et al., (2002), and Safavian and Landgrebe, D. (1991) respectilvey.

**Figure 2.**
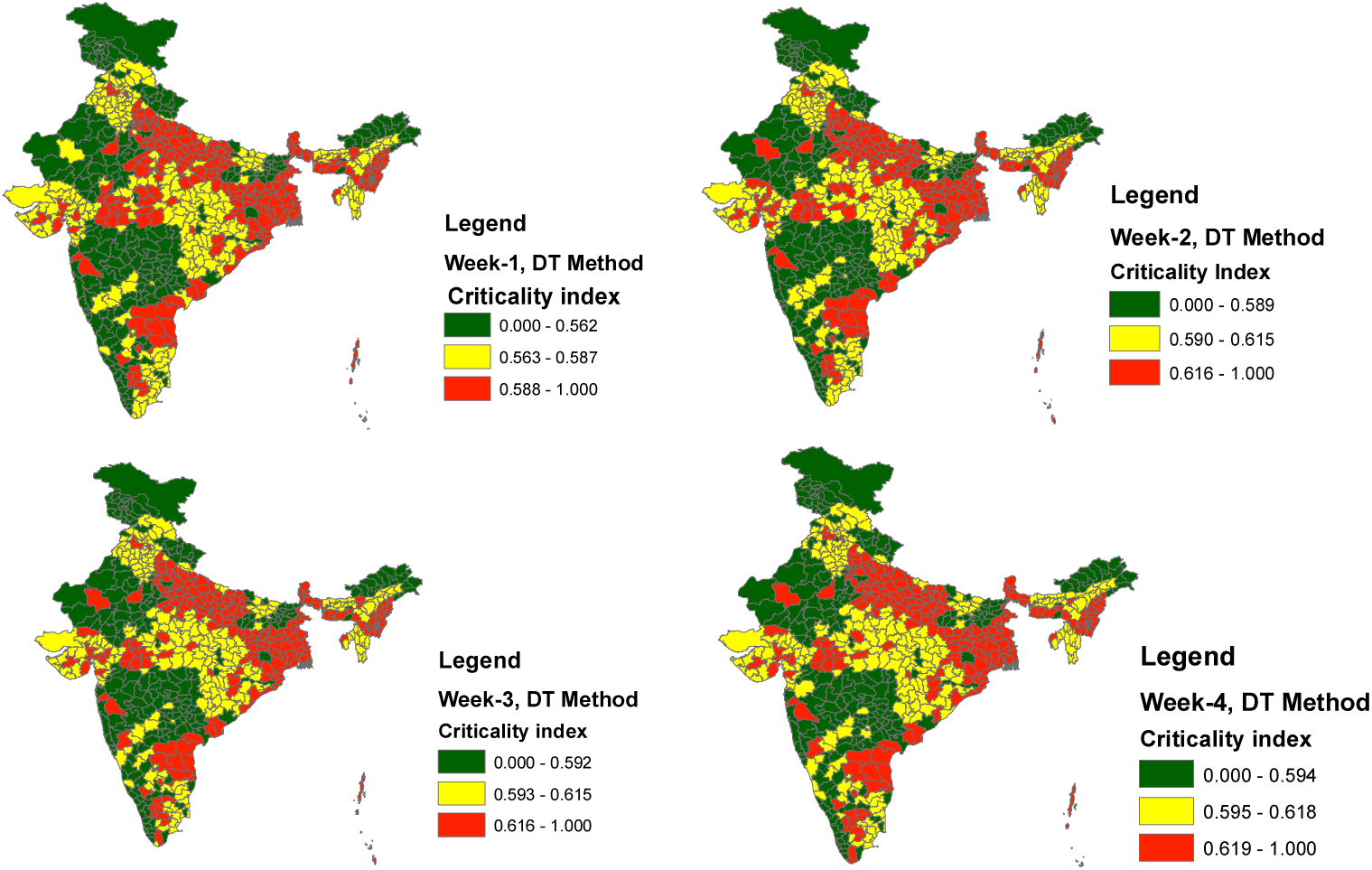
Forecasted risk in terms of criticality index for Week-1, Week-2, Week-3 and Week-4 from 19^th^ April using Decision Tree algorithm

**Figure 3.**
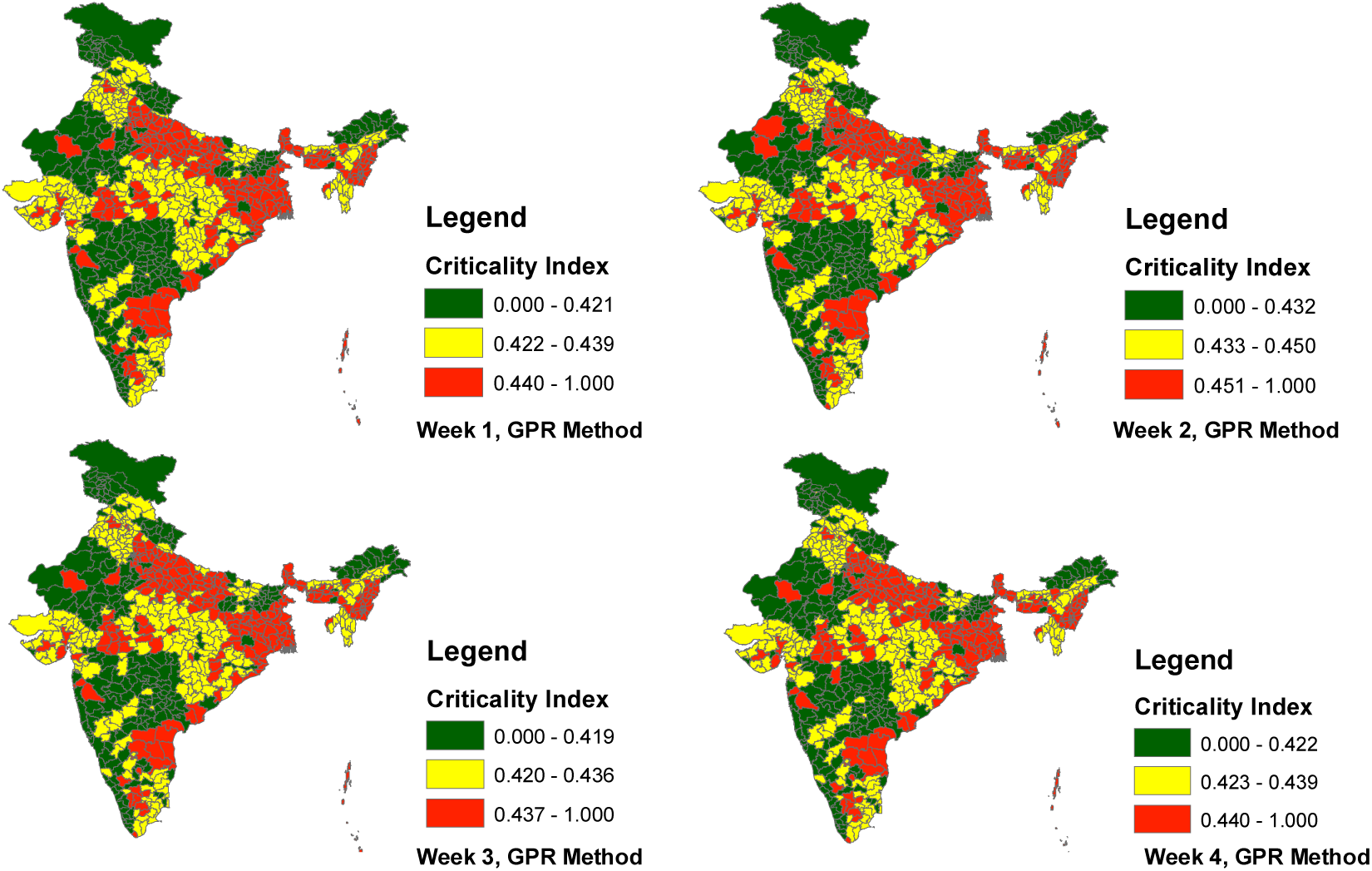
Forecasted risk in terms of criticality index for Week-1, Week-2, Week-3 and Week-4 from 19^th^ April using Gaussian Process Regression

**Figure 4.**
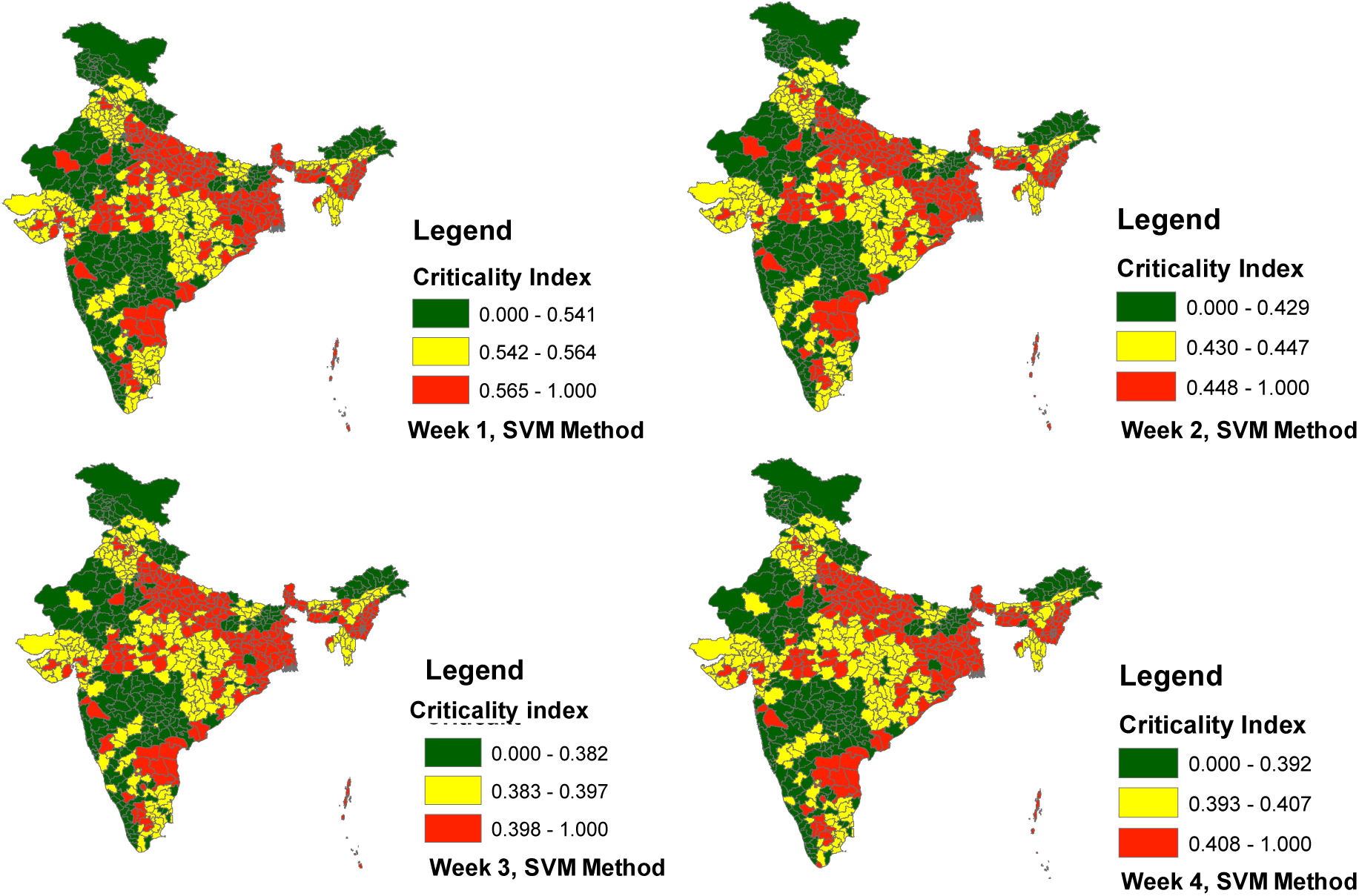
Forecasted risk in terms of criticality index for Week-1, Week-2, Week-3, and Week-4 from 19^th^ April using Support Vector Machine algorithm

It is recommended that districts that are in the green zone can be completely released. However, travel to another adjacent district can be restricted if they fall in another zone. Lockdown can alternatively be opened and closed in these regions with continuous monitoring of the new positive cases.

The importance of the work lies in identifying the districts which are falling in the more severe zone in the following weeks. For such districts, the policy of partial release is recommended with various precautionary actions in place. Due to the nature of the problem, it is recommended that maps should be updated daily, and change of district from one critical zone to another should be identified. Additionally, it is recommended to analyze the decision to identify hotspots as a Multicriteria problem rather than making the decision on a single parameter like the number of cases in a region. For a better understanding of this fact, the statistics of a few of the states are shown in Table 1, and the identified limitations are put as remarks. Therefore, it can conclusively be stated that decisions just based on the number of cases may lead to inadmissible strategies.

From the attribute mentioned in Table 1, it is apparent that apart from active COVID cases, other factors also play an important role, and that is why deceased cases are different. The difference can be attributed to medical infrastructure, average age/ gender of citizens in the district, literacy rate, socio-culture issues, and administration

**Table 1.**
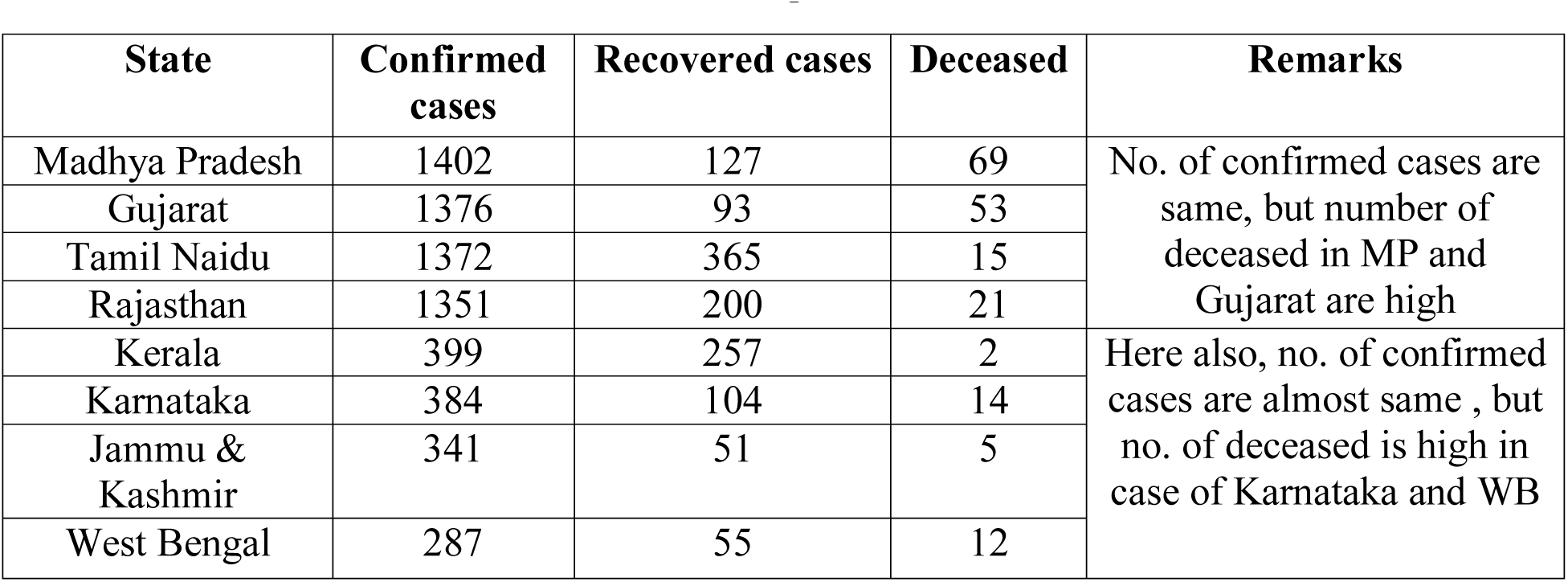
Observed cases, Recovered cases and Deceased cases for a few of the regions as of 19^th^ April

From the mapped risk in terms of the criticality index, it is observed that for a few of the districts, the trend is going to come down, whereas, for a few of the districts, it is anticipated that the cases would rise. It may have to be noted since, for the majority of the period, there is a significant rise in trend of the positive cases in most of the region, it is quite natural that the historical characteristics of the curve will be reflected in the predictions made. Owing to this nature, there are possible chances of reality deviate with the prediction of the long term. In this regard, it is recommended to update the repository and revise the forecast maps on a daily/weekly basis. This exercise would certainly help to combat the COVID-19.

In this study machine, learning-based algorithms are proposed to observe the transmission pattern of Covid-19. Performance evaluation of the forecast is estimated in terms of Root Mean Square Error(RMSE) and coefficient of determination, and the statistics are found to be within satisfactory limits. The model architecture is optimized using Exponential GPR, quadratic SVM, and Fine decision tree out of which, model optimized using exponential GPR showed the best performance with the least RMSE and highest coefficient of determination values. Though the findings help to plan the combat strategies, updating the database would help to forecast which could mimic the reality. The following are a few of the assumptions in the current study that may test the capabilities of the decision-maker in planning the essential strategies since there always be possible for a certain deviation. For better clarity, the assumptions are listed below

Assumption-1: For district-wise analysis, the dataset used in this study for forecasting was taken from various sources^1^. Due to the sudden mass outbreak of this pandemic, there is a discrepancy in data over various sources.

Assumption-2: It may have to be noted that the time series analysis for the total number of daily cases is considered from the date when the first infection was registered in the state. Also, those days were considered as zero values when no case appeared in the district and vice versa for recoveries and deaths for all the districts, respectively.

Assumption-3: For missing data for the district, proportionate value to the nearest hotspot in the state is taken. In forecasting, if the daily increase in the number of cases is less than 0.4, then we have considered it as zero, and if the daily increase in the number of cases is greater than 0.4, then we have considered it as one.

Assumption-4: District-wise deceased cases and recovered cases are based on the new cases which arose 15 days before.

## 4. Conclusion

This study discussed the possible attributes that lead to the formation of a hotspot. Considering the historic data related daily positive new cases, population, population density, deceased cases, availability of hospitals, no of tests conducted, recovered cases, and availability door delivery services, the risk associated with the region is evaluated. Besides evaluating the present, efforts are made to project to the attributes using Machine Learning tools to forecast the possible variation in the discussed parameters. Three different techniques, such as Decision Tree algorithm, Support Vector Machine algorithm, and Gaussian Process Regression, are used to project the data and capture the possible deviation. Additionally, this study proposed a criticality index that can help to quantify the risk in a region which is further used to classify the regions into zones of high risk, low risk, and moderate risk. Developing maps by considering the updated data and mapping the risk for the following weeks by integrating machine learning tools and mapping would certainly help to combat the COVID-19 transmission.

## Data Availability

The authors confirm that the data supporting the findings of this study are available within the article [and/or] its supplementary materials. However it may be noted that the data used for simulations, are taken from the available online data repositories and the same is acknowldged.

## Footnotes

1 https://www.mohfw.gov.in/. https://www.covidl9india.org/

